# Interventions to Reduce Pediatric Cancer Treatment Abandonment in Low- and Middle-Income Countries: A Scoping Review

**DOI:** 10.1101/2025.08.29.25334743

**Authors:** Srinithya R. Gillipelli, Archer R. Schaeffer, Casey McAtee, Constance Nyasulu, Samuel Makuti, R Naitala Andrew, Heather A. Haq, Mark C. Zobeck

## Abstract

**Background:** Pediatric cancer treatment outcomes in low- and middle-income countries (LMICs) significantly lag behind those in high-income countries. One reason for this disparity is high treatment abandonment rates, defined as a failure to start or complete curative-intent therapy after a cancer diagnosis. We conducted a scoping review to describe interventions aimed at reducing treatment abandonment in pediatric cancer patients in LMICs.

**Methods:** Studies identified through systematic database search were included if they met the following criteria: (a) studied an intervention on treatment abandonment; (b) included cancer patients ≤18 years of age; (c) conducted in LMICs as defined by the World Bank income classification; (d) contained pre- and post-intervention measures of treatment abandonment. We restricted inclusion to English-language publications. Two reviewers independently screened the eligible publications and extracted the data. Interventions were categorized as focusing on socioeconomic support, education/psychosocial support, or clinical care quality and capacity improvements (e.g., care navigation, coordination, or therapeutic/diagnostic expansion).

**Findings:** Among the 1,808 articles identified in the search, 21 studies met inclusion criteria: four from the WHO African region, nine from the Americas, seven from the South-East Asian, and three from the Western Pacific Region. Sixteen studies (66%) focused on one category for improvement, and eight (34%) were a mixture of two or more categories. All studies demonstrated a decrease in treatment abandonment after the intervention. The median absolute risk reduction (ARR) was 16% (interquartile range [IQR] 10%-24%). The median relative risk reduction was 72% (IQR 60%-82%). The median pre-intervention abandonment rate across full-text studies was 27% (IQR 20%–34%) and decreased to 7% (IQR 3%–12%) after intervention. Interventions with the largest ARR values included components of socioeconomic support, psychosocial support, and clinic care improvements. All of the 10 studies reporting pre- and post-intervention survival outcomes reported increases in survival following the intervention.

**Interpretation:** Our scoping review describes interventions that were associated with reduced pediatric cancer treatment abandonment in LMICs. Interventions that combined socioeconomic support, psychosocial support, and clinical care quality/capacity improvements yielded the largest reductions. Despite these encouraging findings, limitations of the evidence, including short study durations, single-center designs, lack of control groups, and likely publication bias, restrict the generalizability of results. These findings suggest that treatment abandonment is a targetable and potentially modifiable challenge in LMICs, and that survival outcomes can improve when health systems adopt multifaceted interventions that support families and strengthen care delivery.

**Funding:** National Institutes of Health, K12CA090433.

## Introduction

### Background

Pediatric cancer treatment outcomes in low- and middle-income countries (LMICs) significantly lag behind those in high-income countries. Whereas cure rates for pediatric cancers may reach as high as 90% in high-income countries, cure rates range from 5-60% in LMICs.^1–3^ Moreover, four out of every five cases of childhood cancer occur in LMICs.^4^ Pediatric cancer treatment abandonment, defined as a failure to start or complete curative therapy after a cancer diagnosis, is one of the major drivers of these disparities.^5,6^ These high abandonment rates represent an important target to improve outcomes.

A large amount of literature exists that describes the reasons for treatment abandonment and its clinical implications.^5–7^ Broadly, lack of financial resources, medical facilities, and social support services contribute to the treatment abandonment rates we see in LMICs. Each of these barriers to treatment intersects and exacerbates the others. Decreased access to medical facilities makes travel more costly and burdensome. As the financial and social costs accrue throughout treatment, greater strain is placed on families with fewer social support services.^8^ At medical institutions where poorer patients may receive less individualized attention from oncologists, parents may not be adequately educated on the importance of continuing their child’s therapy.^9^ A poor prognosis or the prospect of a radical or mutilating surgery may trigger treatment abandonment.^10^ In some cases, families have also discontinued therapy when the child demonstrated a strong early response to treatment.^11^

While much is known about the reasons for treatment abandonment, there has been no systematic evaluation of interventions to address this problem. Many interventions have been reported using different methods with different outcome measures across varied treatment contexts. The existing research has major limitations: most studies are single-center, short-term, and use uncontrolled designs, which limits generalizability and sustainability of findings. To synthesize and condense this literature, we chose a scoping review methodology because it enables the mapping of heterogeneous evidence, identification of knowledge gaps, and clarification of key concepts across diverse study designs. We conducted a scoping review to describe interventions that aim to reduce treatment abandonment in pediatric cancer patients in LMICs and to evaluate their impact on treatment abandonment outcomes.

## Methods

### Search Strategy

We followed the Preferred Reporting Items for Systematic Reviews and Meta-Analyses (PRISMA) 2020 guidelines to conduct the literature review.^12^ A comprehensive search was conducted on October 21, 2022, across multiple databases, including MEDLINE Ovid, Embase, Web of Science, CINAHL, and Cochrane. A medical librarian with expertise in systematic reviews developed the initial Medline Ovid search strategy, which we refined using MeSH headings and synonymous keywords/phrases. The strategy was first developed in Medline Ovid using MeSH headings and synonymous keywords/phrases, including the names of LMIC economies according to the World Bank list of countries and lending groups.^13^ The other concepts included Pediatrics, Neoplasms, Treatment Refusal, and intervention types such as Patient Education, Counseling, Self-Help Groups, Patient Advocacy, Patient Navigation, and Patient Identification Systems (Appendix A). Once finalized, the strategy was translated to Embase, Web of Science, CINAHL, and Cochrane, resulting in a total of 2,814 results before and 1,808 results after deduplication (Figure 1).

**Figure 1.**
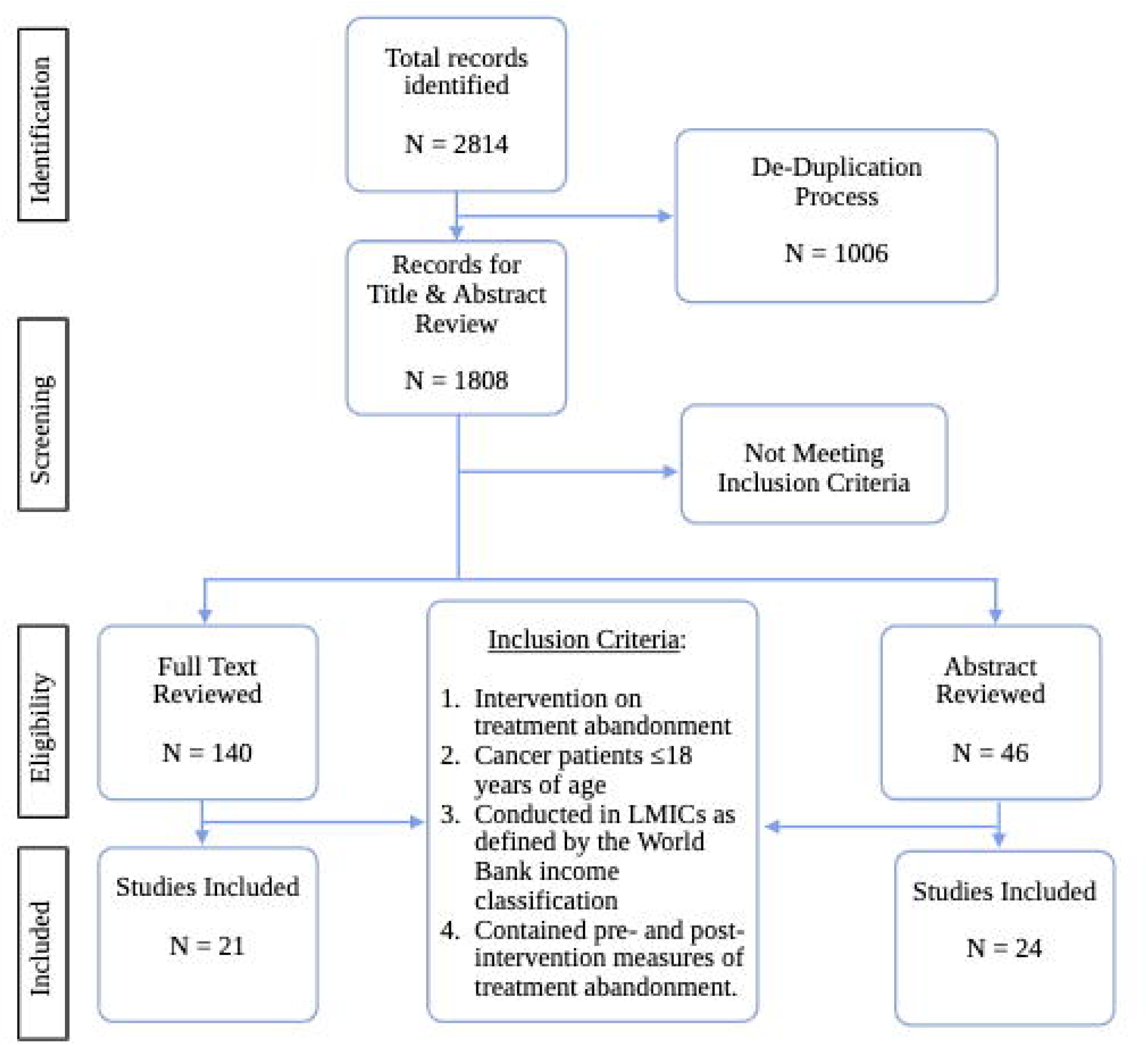
Flow Diagram of Study Selection. This chart depicts the process of screening and selecting studies to include in this review, as well as primary reasons for exclusion among full-text articles.

### Screening and Selection

The review protocol used was based on recommended methods first proposed by Arskey and O’Malley and further developed by Levac et al.^14^ The steps in the review protocol are available in Appendix B. Studies were included if they met the following inclusion criteria: (a) studied an intervention on treatment abandonment; (b) included cancer patients ≤18 years of age; (c) conducted in LMICs as defined by the World Bank income classification; (d) contained pre- and post-intervention measures of treatment abandonment. We restricted the search to studies published in English, which may have excluded relevant studies in other languages. Two authors independently screened the eligible titles and abstracts to identify a total of 140 publications for full-text review. The same two authors independently performed the full-text reviews and identified 21 publications eligible for inclusion in the final analysis (Figure 1). When inclusion was unclear, a third reviewer resolved the decision.

### Categorizing Interventions

The interventions observed in the final selection varied widely. These interventions involved, but were not limited to, providing fully funded healthcare through private donation, hospital twinning programs, adding social workers to the care team, reaching out to patients after loss to follow-up, and subsidizing transportation fees. These interventions were classified into three categories: socioeconomic support, education/psychosocial support, and clinical care improvements (defined as interventions that enhance diagnostic and therapeutic capacity, care navigation/coordination, or overall quality of clinical services). The study structure was classified either as 1) a prospective, treatment abandonment-specific intervention with a concurrent control, 2) a prospective treatment abandonment-specific intervention with a pre-post design, 3) a general (not directly targeting abandonment) prospective intervention with a pre-post design, and 4) a retrospective pre-post design.

### Data extraction

Two reviewers independently screened the eligible publications and extracted the data, including title, publication year, journal of publication, single or multi-site, country of study, study design and population, components of intervention, survival results, and abandonment results. To ensure the consistency of the extracted data elements between reviewers, both reviewers extracted data separately and sequentially for a common test set of 10 articles. The results were compared for consistency and discussed between the two reviewers and the research team. By the end of the 10-article test set, data extraction was consistent across reviewers. Data were then extracted from the remaining articles and abstracts. Any questions that arose during data extraction were discussed between the reviewers and triangulated with a third reviewer.

### Data synthesis

We summarized the extracted data across studies and generated descriptive statistics. We calculated the absolute risk reduction (ARR) as the pre-intervention abandonment rate minus the post-intervention abandonment rate, representing the absolute decrease in treatment abandonment. We calculated the relative risk reduction (RRR) as one minus the ratio of the post-intervention abandonment rate to the pre-intervention abandonment rate. Analyses were stratified by abstract-only reports and full-text articles.

## RESULTS

### Full-text Articles

Twenty-one full studies met the eligibility criteria. These studies were distributed across different regions, with four (19%) from the WHO African region, seven (33%) from the Americas, seven (33%) from the South-East Asian region, and three (14%) from the Western Pacific Region. The studies employed various methodologies to assess interventions, with six (29%) using treatment abandonment-specific prospective pre-post studies, ten (48%) using retrospective pre-post studies, and five (24%) using general intervention pre-post studies. No TxA-specific intervention studies leveraged a concurrent control to evaluate the intervention. Interventions encompassed a range of components aimed at addressing the multifaceted challenges contributing to treatment abandonment. These components included financial support, material assistance, transportation assistance, education for caregivers and providers, psychosocial support, facility-level improvements, and care navigation/patient-tracking systems. Among the interventions, six (29%) focused exclusively on socioeconomic support, five (24%) on improving clinical care, and three (14%) on improving education/psychosocial support. Additionally, four (20%) were a mixture of two categories, and three (14%) included components from all three categories. Study demographics are described in Table 1.

**Table 1.**
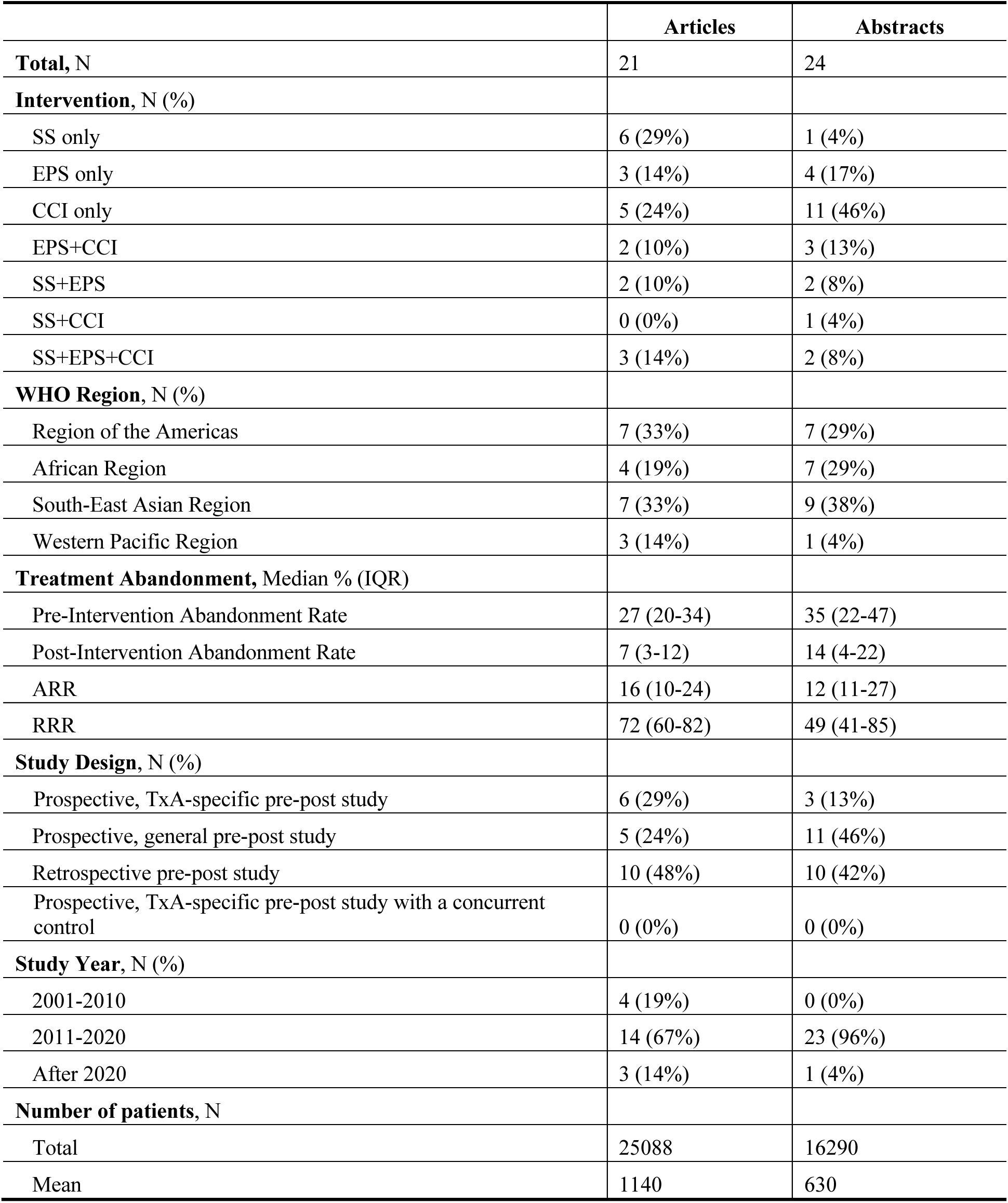
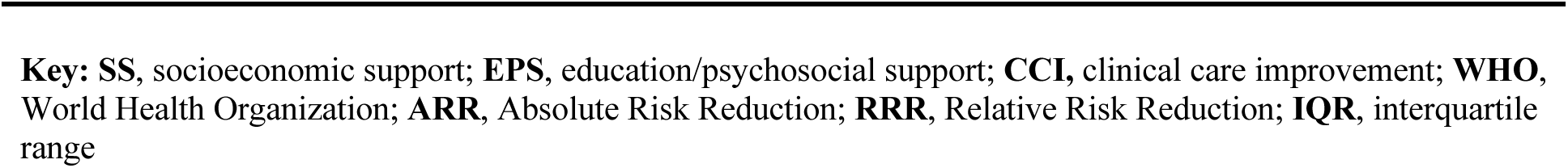
Descriptive Summary Characteristics of Included Studies.

|  | Articles | Abstracts |
| --- | --- | --- |
| **Total, N** | 21 | 24 |
| **Intervention, N (%)** |  |  |
| SS only | 6 (29%) | 1 (4%) |
| EPS only | 3 (14%) | 4 (17%) |
| CCI only | 5 (24%) | 11 (46%) |
| EPS+CCI | 2 (10%) | 3 (13%) |
| SS+EPS | 2 (10%) | 2 (8%) |
| SS+CCI | 0 (0%) | 1 (4%) |
| SS+EPS+CCI | 3 (14%) | 2 (8%) |
| **WHO Region, N (%)** |  |  |
| Region of the Americas | 7 (33%) | 7 (29%) |
| African Region | 4 (19%) | 7 (29%) |
| South-East Asian Region | 7 (33%) | 9 (38%) |
| Western Pacific Region | 3 (14%) | 1 (4%) |
| **Treatment Abandonment, Median % (IQR)** |  |  |
| Pre-Intervention Abandonment Rate | 27 (20-34) | 35 (22-47) |
| Post-Intervention Abandonment Rate | 7 (3-12) | 14 (4-22) |
| ARR | 16 (10-24) | 12 (11-27) |
| RRR | 72 (60-82) | 49 (41-85) |
| **Study Design, N (%)** |  |  |
| Prospective, TxA-specific pre-post study | 6 (29%) | 3 (13%) |
| Prospective, general pre-post study | 5 (24%) | 11 (46%) |
| Retrospective pre-post study | 10 (48%) | 10 (42%) |
| Prospective, TxA-specific pre-post study with a concurrent control | 0 (0%) | 0 (0%) |
| **Study Year, N (%)** |  |  |
| 2001-2010 | 4 (19%) | 0 (0%) |
| 2011-2020 | 14 (67%) | 23 (96%) |
| After 2020 | 3 (14%) | 1 (4%) |
| **Number of patients, N** |  |  |
| Total | 25088 | 16290 |
| Mean | 1140 | 630 |

**Key:** **SS**, socioeconomic support; **EPS**, education/psychosocial support; **CCI**, clinical care improvement; **WHO**, World Health Organization; **ARR**, Absolute Risk Reduction; **RRR**, Relative Risk Reduction; **IQR**, interquartile range

The interventions varied in terms of the specific components included, duration of implementation, and delivery methods (Table 2). Some interventions provided ongoing support throughout the course of treatment, while others were targeted interventions delivered over a defined period. The studies reported treatment abandonment rates both before and after the intervention, allowing for a comprehensive analysis of the effectiveness of the interventions. All studies demonstrated a decrease in treatment abandonment after the intervention. Data are reported as median and interquartile range (IQR). The median pre-intervention abandonment rate was 27% (IQR 20%–34%) and decreased to 7% (IQR 3%– 12%) after intervention. The median ARR was 16%, with an IQR of 10% to 24%, while the median RRR was 72% with an IQR of 60% to 82%. Comparing interventions across categories revealed that interventions combining socioeconomic support, psychosocial support, and clinical care improvements showed the largest reductions in treatment abandonment rates. All of the 10 studies reporting pre- and post-intervention survival outcomes reported increases in survival following the intervention.

**Table 2.** Manuscripts Evaluating Treatment Abandonment Interventions.

| Category | Article Reference | Location | WHO Region | Timeframe | Study design | Target Disease | Number of patients | Pre- TxA Rate (%) | Post-TxA Rate (%) | ARR (%) | RRR (%) | Survival Outcomes |
| --- | --- | --- | --- | --- | --- | --- | --- | --- | --- | --- | --- | --- |
| CCI | Israels, 2018 (Pediatr Blood Cancer) | Cameroon, Ghana, Malawi | Africa | 2014/01/01 to 2016/01/01 | Prospective pre-post study | Wilms Tumor | 122 | 23 | 13 | 10 | 43 | 52% to 68%, 2y “end of treatment period” outcome |
| CCI | Chagaluka, 2020 (Pediatr Blood Cancer) | Malawi, Cameroon, Ghana | Africa | 2014/01/01 to 2018/01/01 | Prospective pre-post study | Wilms Tumor | 201 | 23 | 12 | 11 | 48 | 52% 1yr to 68.5% 3y “end of treatment period” outcome |
| SS | Morgan, 2022 (Pediatr Blood Cancer) | Tanzania | Africa | 2017/05/01 to 2018/04/01 | Retrospective pre-post study | Any cancer | 149 | 37 | 27 | 10 | 27 | From 28% 1yr EFS to 34% 1yr EFS |
| SS | Chakumatha, 2022 (Pediatr Blood Cancer) | Malawi | Africa | 2020/06/01 to 2019/06/01 | TxA-specific prospective pre-post study | ALL, HD, Wilms, Rb, BL | 414 | 19 | 7 | 12 | 63 | 53% to 61% |
| CCI | Howard, 2004 (JAMA) | Brazil | Americas | 1980/01/01 to 2002/01/01 | Retrospective pre-post study | ALL | 375 | 16 | 0.5 | 15.5 | 97 | 5 Year EFS 32% over 9yrs to 63% over 5yrs |
| EPS | Ferman, 2019 (Pediatr Blood Cancer) | Brazil | Americas | 2012/01/01 to 2017/12/31 | Retrospective pre-post study | Solid tumors | 1139 | 2.5 | 0.9 | 1.6 | 64 | 47.4% (no baseline) |
| EPS, CCI | Wilimas, 2010 (Pediatr Blood Cancer) | Guatemala, Honduras, El Salvador | Americas | 2000/01/01 to 2008/01/01 | Prospective pre-post study | Rb | 363 | 21 | 11 | 10 | 48 | 40% 3yr Survival to 61% 4 yr survival |
| EPS, CCI | Suarez, 2015 (Pediatr Blood Cancer) | Colombia | Americas | 2007/08/31 to 2010/01/01 | TxA-specific prospective pre-post study | ALL | 280 | 32 | 9 | 23 | 72 | 47% 2yr EFS to 65% 2yr EFS |
| SS, EPS | Alvarez, 2017 (Pediatr Blood Cancer) | Guatemala | Americas | 2001/01/01 to 2008/8/9 | Retrospective pre-post study | Any cancer | 1789 | 27 | 7 | 20 | 74 | 57% over 8yrs, no baseline |
| SS, EPS, CCI | Luna-Fineman, 2019 (J Clin Oncol) | Guatemala, Honduras, El Salvador, Nicaragua | Americas | 2007/04/01 to 2015/12/01 | Prospective pre-post study | Rb | 161 | 20 | 4 | 16 | 80 | From 70% 5yr EFS to 81% 5 yr EFS |
| SS, EPS, CCI | Salaverria, 2015 (Pediatr Blood Cancer) | El Salvador | Americas | 2011/01/01 to 2012/12/01 | TxA-specific prospective pre-post study | Any cancer | 491 | 13 | 3 | 10 | 77 | N/A |
| EPS | Mostert, 2009 (Arch Dis Child) | Indonesia | South-East Asia | 2004/01/01 to 2006/12/01 | Retrospective pre-post study | ALL | 283 | 34 | 26 | 8 | 24 | 24% 2yr EFS to 31% 2yr EFS |
| EPS | Kumar, 2013 (Pediatrc Blood Cancer) | India | South-East Asia | 2008/03/01 to 2011/08/01 | Retrospective pre-post study | Rb | 101 | 71.4 | 16.7 | 54.7 | 77 | N/A |
| SS | Srinivasan, 2015 (Indian J Palliat Care) | India | South-East Asia | 2010/01/01 to 2013/06/01 | Retrospective pre-post study | Any cancer | 170 | 1.4 | 0 | 1.4 | 100 | 80% (no baseline) |
| SS | Jatia, 2019 (Pediatr Blood Cancer) | India | South-East Asia | 2009/01/01 to 2016/01/01 | TxA-specific prospective pre-post study | Any cancer | 11011 | 20 | 3.7 | 16.3 | 82 | N/A |
| SS | Alam, 2019 (Psychooncology) | India | South-East Asia | 1995/01/01 to 2017/12/31 | Retrospective pre-post study | ALL | 1041 | 30.2 | 11.1 | 19.1 | 63 | N/A |
| SS, EPS | Moulik, 2016 (J Pediatr Hematol Oncol) | India | South-East Asia | 2007/01/01 to 2013/09/01 | TxA-specific prospective pre-post study | ALL | 418 | 33.4 | 9.3 | 24.1 | 72 | N/A |
| SS, EPS, CCI | Narula, 2018 (Indian J. Cancer) | India | South-East Asia | 2010/01/01 to 2016/12/01 | TxA-specific prospective pre-post study | Hematolymphoid cancers | 5261 | 32 | 3.4 | 28.6 | 89 | N/A |
| CCI | Luo, 2009 (Pediatr Blood Cancer) | China | Western Pacific | 1999/12/01 to 2004/09/01 | Prospective pre-post study | Acute Promyelocytic Leukemia | 30 | 37.5 | 0 | 37.5 | 100 | 37.5% 3.5yr EFS to 79.6% 3.5 yr EFS |
| CCI | Gaston, 2021 (Asian Pac J Cancer Prev) | Philippines | Western Pacific | 2016/01/01 to 2019/6/30 | Retrospective pre-post study | Osteosarcoma | 75 | 50 | 6 | 44 | 88 | N/A |
| SS | Zhou, 2015 (J Pediatr Hematol Oncol) | China | Western Pacific | 2002/01/01 to 2012/12/01 | Retrospective pre-post study | ALL | 1151 | 50 | 20 | 30 | 60 | N/A |

Key: **TxA**, treatment abandonment; **CCI**, Clinical Care Improvement; **SS**, Socioeconomic Support; **EPS**, Educational/Psychosocial Support; **EFS**, Event-free survival; **Pre- and Post- TxA Rate**, Pre- and Post- Intervention Treatment Abandonment Rate; **ARR**, Absolute Risk Reduction; **RRR**, Relative Risk Reduction; **ALL**, acute lymphoblastic leukemia; **HD**, Hodgkins disease; **Rb**, retinoblastoma; **BL**, Burkitt lymphoma.

### Education/Psychosocial Support

The three interventions focused only on education and psychosocial support aimed to improve caregiver understanding, provide counseling, and address sociocultural factors that contribute to treatment abandonment. These interventions targeted both patients and caregivers with structured education, psychosocial counseling, and follow-up support to strengthen adherence. Mostert et al. introduced a structured parental education program in Indonesia targeting parents of children with acute lymphoblastic leukemia. The program included educational materials such as videos, booklets, and DVDs to improve understanding of treatment and access to donated chemotherapy.^15^ Ferman et al. created an intervention in Brazil aimed at preventing treatment abandonment for children with solid tumors by implementing a patient-tracking system and providing early interventions to address issues associated with treatment.^16^ In India, Kumar et al. focused on the implementation of effective pre-abandonment counseling by a support team to address concerns and barriers to treatment adherence, particularly in rural populations.^17^ All three interventions focused on strengthening communication, enhancing caregiver knowledge, and mitigating emotional or social stressors linked to treatment discontinuation. Across these studies, the median ARR was 8% (IQR 5%-31%), and the median RRR was 64% (IQR 44%-70%).

### Socioeconomic Support

The six interventions that focused primarily on socioeconomic support shared common goals of reducing treatment abandonment by addressing financial and logistical barriers to care. In Malawi, Chakumatha et al. implemented a comprehensive model that fully subsidized treatment, transport, accommodation, and food for families, alongside a care navigation system to reduce missed appointments.^18^ In India, Jatia et al. evaluated the impact of holistic financial assistance, including support from non-governmental organizations (NGOs), facility-level improvements, and tracking systems.^19^ Srinivasan et al. focused on direct financial aid from private cancer support groups, demonstrating complete elimination of abandonment in their sample.^20^ Alam and Kumar also evaluated the role of financial support in India and found that economic aid alone was insufficient to reduce abandonment unless combined with counseling.^21^ Their findings highlight that financial interventions may need to be paired with psychosocial components to achieve maximum effect. In China, Zhou et al. examined government-funded healthcare programs and found substantial reductions in abandonment across both urban and rural populations, including a drop from 89% to 3% in rural areas.^22^ Morgan et al. assessed the impact of a residential hostel in Tanzania, providing free accommodation for pediatric cancer patients and their caregivers. The intervention led to a 10% absolute reduction in abandonment and a modest increase in one-year survival, suggesting that housing support can reduce logistical obstacles to consistent care. However, this was a private program that did not cover medications or hospital costs, and abandonment rates remained >25% overall in the setting.^23^ Across these six studies, the median ARR was 14% (IQR 11%-18%), and the median RRR was 63% (IQR 61%-77%), demonstrating the consistent effectiveness of socioeconomic interventions in improving adherence to pediatric cancer treatment.

### Clinical Care Improvement

Five interventions focused on strengthening the quality and delivery of clinical care to reduce treatment abandonment and improve outcomes. Israels et al. conducted a multicountry study of Wilms tumor in Cameroon, Ghana, and Malawi that implemented adapted treatment guidelines alongside basic supportive care improvements, resulting in a 10% absolute reduction in abandonment and a relative improvement in survival outcomes. Chagaluka et al., extended the prior analysis, demonstrating persistent reduction in abandonment and gains in event-free survival among a larger cohort with more follow-up time.^24^ In Brazil, Howard et al. established a dedicated pediatric oncology unit with protocol-based therapy, multidisciplinary care, 24-hour physician coverage, and provider training. This intervention led to a dramatic reduction in abandonment from 16% to 0.5% and an increase in five-year event-free survival from 32% to 63%.^25^

In China, Luo et al. compared two treatment regimens for childhood acute promyelocytic leukemia (APL), finding that a reduced-intensity protocol based on PETHEMA LPA99 resulted in higher survival, fewer complications, and lower costs..^26^ Gaston et al. implemented a patient navigator program in the Philippines to improve follow-up and reduce abandonment among patients with high-grade osteosarcoma, decreasing abandonment from 50% to 6%.^27^ Together, these studies demonstrate the effectiveness of clinical interventions, ranging from treatment protocol optimization to patient navigation and facility-based care enhancements, in improving pediatric cancer adherence. Clinical care improvement interventions yielded a median ARR of 16% (IQR 11%–38%) and median RRR of 88% (IQR 48%–97%), highlighting their high impact on treatment retention and survival.

### Combined Interventions

Seven studies implemented interventions spanning multiple categories–socioeconomic support (SS), education/psychosocial support (EPS), and clinical care improvement (CCI)-to address the multifactorial barriers contributing to treatment abandonment.

Two studies combined EPS and CCI. Wilimas et al. implemented provider education, standardized treatment protocols, and facility-level improvements in Central America, aiming to strengthen local capacity and improve care delivery. These were paired with educational outreach to enhance caregiver understanding.^28^ Suarez et al. in Colombia used an approach that integrated risk-adapted treatment regimens with psychosocial support and provider training to reduce abandonment and mortality.^29^

Two studies combined SS and EPS: Alvarez et al. in Guatemala developed a comprehensive program that included financial, material, and transportation support alongside education for caregivers and providers, psychosocial services, and a patient tracking system.^30^ Moulik et al. in India provided financial subsidies for treatment along with structured, multistage counseling sessions to address parental concerns and promote adherence.^31^

Three studies incorporated all three components (SS + EPS + CCI): Luna-Fineman et al. implemented a neoadjuvant chemotherapy protocol to address enucleation refusal in retinoblastoma, combined with psychosocial support and socioeconomic assistance across multiple Latin American countries.^32^ Salaverria et al. in El Salvador introduced financial support, caregiver education, and a time-sensitive adherence tracking system to improve follow-up and reduce missed appointments.^33^ Narula et al. in India implemented a wide-ranging intervention involving financial/material aid, caregiver and provider education, psychosocial services, and treatment/facility-level improvements.^34^

All of these interventions share the goal of improving treatment adherence, but differ in their specific strategies, which include early diagnosis, psychosocial support, tracking procedures, reduced-intensity treatment regimens, and neoadjuvant chemotherapy. The two studies combining both clinical care improvement and educational and psychosocial support (Wilimas 2010 and Suarez 2015) had a median ARR of 17% and median RRR of 60%.^28,29^ The two studies combining both socioeconomic support and educational and psychosocial support (Alvarez 2017 and Moulik 2016) had a median ARR of 22% and median RRR of 73%.^30,31^ The three studies focusing on all three intervention components (Luna-Fineman 2019, Salaverria 2015, and Narula 2018) had a median ARR of 16% and median RRR of 80%.^32–34^ These results suggest that combining multiple forms of support may yield greater reductions in treatment abandonment. In particular, multifaceted interventions that include socioeconomic, clinical, and psychosocial components appear to be the most effective strategies for improving continuity of care in pediatric oncology across low- and middle-income countries.

### Abstracts

A total of 24 abstracts met eligibility criteria after removing duplicates and those with corresponding full-text articles already included (Table 3). These studies span more than 10 countries across the WHO African Region, South-East Asian Region, Western Pacific Region, and Region of the Americas, and all were conducted in lower- or upper-middle income countries. The majority used retrospective or prospective pre-post designs. The median pre-intervention abandonment rate was 35% (IQR 22%–47%) and decreased to 14% post-intervention (IQR 4%–22%). The median ARR was 12% and the median RRR was 49%. Although improvements were observed, post-intervention abandonment rates of ∼14% remain substantially higher than those in wealthier LMICs (<5%) and nearly absent in high-income countries. The most substantial reductions were observed in studies implementing multicomponent interventions, while programs focused on a single intervention tended to show more modest improvements.^35–48^

**Table 3.** Abstracts Evaluating Treatment Abandonment Interventions.

| Category | Article Reference | Location | WHO Region | Timeframe | Study design | Number of patients | Pre- TxA Rate (%) | Post- TxA Rate (%) | ARR (%) | RRR (%) | Survival Outcomes |
| --- | --- | --- | --- | --- | --- | --- | --- | --- | --- | --- | --- |
| CCI, EPS | Achempim, 2016 (Pediatri Blood Cancer) | Ghana | African Region | 2010/01/01 to 2015/01/01 | Prospective pre-post study | ---- | 40 | 11.3 | 28.7 | 72 | ---- |
| EPS, SS | Banerjee, 2018 (Pediatric Hematology Oncology Journal) | India | Southeast Asian Region | 2011/05/01 to 2018/01/01 | Retrospective pre-post study | 2223 | 1.8 | 0.05 | 1.75 | 97 | ---- |
| CCI, EPS, SS | Bhattacharya 2019 (Pediatri Hematology Oncology) | India | Southeast Asian Region | 2013/01/01 to 2018/01/01 | Prospective pre-post study | 2015 | 15.3 | 8.3 | 7 | 46 | 51.9% to 75.3% |
| SS | Cruz, 2018 (Pediatri Blood and Cancer) | Philippines | Western Pacific Region | 2013/01/01 to 2017/01/01 | Prospective pre-post study | 470 | 14.4 | 3.8 | 10.6 | 74 | EFS: 92.4% to 93.9%<br>OS: 84.3% to 91.2% |
| CCI | Day, 2015 (Pediatri Blood and Cancer) | Central America | Region of the Americas | ---- | Prospective pre-post study | 1936 | 10.2 | 6.5 | 3.7 | 36 | ---- |
| EPS, CCI | Figueredo, 2013 (Pediatri Blood and Cancer) | Paraguay | Region of the Americas | 2009/01/01 to 2012/08/01 | Prospective pre-post study | 404 | 2.6 | 1.5 | 1.1 | 42 | ---- |
| EPS | Fuentes-Alabi, 2018 (Pediatri Blood and Cancer) | El Salvador | Region of the Americas | 2011/01/01 to 2018/01/01 | TxA-specific prospective pre-post study | --- | 13 | 2 | 11 | 85 | ---- |
| CCI | Gunasekera, 2018 (Pediatri Blood and Cancer) | Sri Lanka | Southeast Asian Region | 2003/01/01 to 2014/01/01 | Prospective pre-post study | 76 | 27 | 15 | 12 | 44 | ---- |
| CCI | Gupta, 2016 (Pediatri Blood and Cancer) | India | Southeast Asian Region | 2015/03/01 to 2016/03/01 | TxA-specific prospective pre-post study | ---- | 50 | 10 | 40 | 80 | 30% to 10% |
| EPS | Igenge, 2021 (Cancer Epidemiology Biomarkers and Prevention) | Tanzania | African Region | 2016/01/01 to 2019/01/01 | Retrospective pre-post study | 55 | 38.2 | 26.7 | 11.5 | 30 | ---- |
| EPS, SS | Jatia, 2012 (Pediatri Blood and Cancer) | India | Southeast Asian Region | 2010/01/01 to 2011/12/01 | TxA-specific prospective pre-post study | 2353 | 10.4 | 5.3 | 5.1 | 49 | ---- |
| CCI, SS | Langat, 2019 (Journal of Clinical Oncology) | Kenya | African Region | ---- | Prospective pre-post study | 106 | 35 | 5 | 30 | 86 | 1-year OS: 29% to 50% |
| CCI | Lassaletta, 2013 (Pediatri Blood and Cancer) | Paraguay | Region of the Americas | 2000/01/01 to 2012/01/01 | Retrospective pre-post study | 587 | 51 | 29 | 22 | 43 | 3yr EFS: 40.5% to 60.7%<br>3yr OS: 57.6% to 68.6% |
| EPS | Libes, 2014 (Pediatri Blood and Cancer) | Kenya | African Region | 2008/01/01 to 2012/12/01 | Prospective pre-post study | 82 | 39.3 | 3.8 | 35.5 | 90 | ---- |
| EPS | Mafwimbo, 2017 (Supportive Care in Cancer) | Tanzania | African Region | 2010/01/01 to 2016/01/01 | Retrospective pre-post study | 305 | 44 | 32 | 12 | 27 | 18% to 29% |
| CCI | Mahajan, 2012 (Pediatri Blood and Cancer) | India | Southeast Asian Region | 2010/01/01 to 2012/01/01 | Retrospective pre-post study | ---- | 60 | 20 | 40 | 67 | ---- |
| CCI | Mehta, 2013 (Pediatri Blood and Cancer) | India | Southeast Asian Region | 2005/01/01 to 2011/01/01 | Retrospective pre-post study | 121 | 60.6 | 24.1 | 36.5 | 60 | 20% to 51.85% |
| CCI | Paintsil, 2020 (Pediatri Blood and Cancer) | Sub-Saharan Africa | African Region | 2014/01/01 to 2020/01/01 | Prospective pre-post study | 201 | 23 | 12 | 11 | 48 | 2yr OS: 52% to 68.5% |
| CCI | Quintero, 2012 (Pediatri Blood and Cancer) | Colombia | Region of the Americas | 1994/01/01 to 2010/01/01 | Retrospective pre-post study | 292 | 35 | 18 | 17 | 49 | EFS: 43% to 46%<br>OS: 55% to 59% |
| CCI, EPS | Samudio, 2018 (Journal of Global Oncology) | Paraguay | Region of the Americas | 2009/01/01 to 2017/12/01 | Prospective pre-post study | 396 | 17.5 | 0 | 17.5 | 100 | ---- |
| CCI | Tagoe, 2018 (Pediatri Blood and Cancer) | Ghana | African Region | 2006/01/01 to 2015/12/01 | Retrospective pre-post study | 604 | 50 | 42 | 8 | 16 | ---- |
| CCI | Tandon, 2015 (Pediatri Blood and Cancer) | India | Southeast Asian Region | 2009/01/01 to 2014/08/01 | Retrospective pre-post study | 188 | 26 | 2.6 | 23.4 | 90 | ---- |
| CCI, EPS, SS | Trehan, 2015 (Pediatri Blood and Cancer) | India | Southeast Asian Region | 2004/01/01 to 2014/06/01 | Retrospective pre-post study | 3568 | 28 | 17 | 11 | 39 | ---- |
| CCI | Valverde, 2012 (Pediatri Blood and Cancer) | Honduras | Region of the Americas | 2000/01/01 to 2011/01/01 | Prospective pre-post study | 138 | 20 | 14 | 6 | 30 | 3yr EFS: 63% to 72% (in those who did not abandon) |

Key: **TxA**, treatment abandonment; **CCI**, Clinical Care Improvement; **SS**, Socioeconomic Support; **EPS**, Educational/Psychosocial Support; **EFS**, Event-free survival; **Pre- and Post- TxA Rate**, Pre- and Post-Intervention Treatment Abandonment Rate; **ARR**, Absolute Risk Reduction; **RRR**, Relative Risk Reduction.

## DISCUSSION

Overall, the results demonstrate that treatment abandonment is a targetable and potentially modifiable problem that can be addressed through dedicated efforts by healthcare providers, national governments, and non-government organizations. These results highlight the effectiveness of interventions that target education, psychosocial support, socioeconomic assistance, and clinical care quality and capacity in reducing treatment abandonment and improving survival. All 21 full-text articles demonstrated a decrease in treatment abandonment rates following the intervention, with notable improvements in survival in all 10 studies reporting outcomes. The findings underscore the significance of targeted interventions in addressing the complex challenges contributing to treatment abandonment in LMICs. Interventions combining socioeconomic support, psychosocial support, and clinical care improvements emerged as particularly effective strategies. Yet no studies used a concurrent control group, most were retrospective or single-center, and the majority evaluated interventions lasting only one to two years, limiting generalizability and sustainability. Despite these improvements, post-intervention abandonment rates remained substantially higher than those reported in upper-middle income countries (<5%) and high-income countries (near zero). The fact that all studies demonstrated improvement further raises concern for publication bias and selective reporting.

The observed reductions in treatment abandonment rates and improvements in survival outcomes have profound implications for the management of pediatric cancer in LMICs. Targeted interventions can potentially enhance patient outcomes and reduce disparities in cancer care. An evaluation of intervention categories revealed varying degrees of effectiveness, with interventions combining multiple components demonstrating the most significant reductions in treatment abandonment rates. However, each category had its strengths and weaknesses, emphasizing the need for multifaceted approaches. Importantly, the available literature is heavily weighted toward upper-middle and lower-middle income countries such as Brazil, China, and India, where economic contexts differ greatly from low-income countries such as Malawi or Tanzania. Interventions that succeed in higher-resource LMICs may not translate to settings with far fewer resources, underscoring a critical gap in the evidence base.

An important component of interventions across all categories was the use of care navigation/patient tracking systems. Pediatric cancer treatment is a process that can take months or years to complete. Treatment centers must have the ability to detect when unplanned treatment absences occur along this process to support patients at risk of abandonment. As many studies in our review have demonstrated the effectiveness of this approach, care navigation systems are a core component of successful strategies to reduce treatment abandonment.

Limitations of the scoping review include potential biases inherent in the included studies, such as selection bias and confounding variables. Notably, no studies that met our inclusion criteria demonstrated ineffective interventions, suggesting that positive publication bias may confound estimates of intervention effectiveness. For this reason, all results should be interpreted as descriptive rather than definitive estimates of effect size. Additionally, the heterogeneity of interventions and outcome measures across studies poses challenges for directly comparing interventions. Most studies were single-center, uncontrolled, and limited in duration, reducing confidence in their generalizability or sustainability.

Finally, it is possible that some relevant studies were excluded due to our restriction to English-language publications. We conducted a thorough literature review to mitigate this risk, and we believe the included literature meets the goal of a scoping review to describe the general body of evidence about interventions to reduce abandonment.

Despite these limitations, the review provides a comprehensive overview of interventions to reduce treatment abandonment in pediatric cancer patients in LMICs. The inclusion of diverse study types across geographic regions enhances the descriptive value of the findings and offers useful insights for practice. Future research should prioritize rigorous evaluations, including cluster-randomized or crossover designs, and adopt standardized definitions of abandonment to allow comparability. Studies in low-income countries are especially needed, with long-term follow-up to evaluate the durability of intervention effects.

Targeted interventions combining socioeconomic support, psychosocial support, and clinical care improvements can reduce treatment abandonment and improve survival outcomes for pediatric cancer patients in LMICs. The findings have implications for clinical practice, policy development, and resource allocation in resource-limited settings. Effective interventions identified in this review can inform the design of targeted interventions tailored to the specific needs of a cancer treatment program.

## Data Availability

All data produced in the present study are available upon reasonable request to the authors

## Acknowledgements

We would like to thank Amy Sisson, MS for her assistance with the literature search.

## Contributors

MZ, HH, and SRG developed the idea (conceptualization). MZ and HH were responsible for funding acquisition and resources. MZ and SRG developed the methodology. MZ, HH, and SRG were responsible for project administration. MZ, SRG, and ARS participated in data collection (data curation). MZ, SRG, and ARS conducted the data analyses (investigation) and had access to the raw data; verified the data. MZ and HH provided supervision. MZ, SRG, and ARS drafted the manuscript and prepared the tables and figures. All authors contributed to the interpretation of the findings, the editing of the article, and the approval of the final submitted version. All authors have access to all the data reported in this study.

## Data Sharing

The protocol, analysis plan, and data for this study will be made available upon reasonable request.

## Declaration of interests and source of funding statements

### Research in context

Evidence before this study: Prior research has highlighted the risk factors for pediatric cancer treatment abandonment in various regions, necessitating interventions to address this issue. Studies have explored interventions targeting factors contributing to treatment abandonment, including socioeconomic barriers, lack of access to healthcare, and inadequate support systems. These studies have provided valuable insights into the effectiveness of different intervention strategies, but a comprehensive analysis of the overall impact and trends across interventions has yet to be conducted.

Added value of this study: We synthesize evidence from multiple studies to provide a comprehensive overview of interventions aimed at reducing treatment abandonment rates in diverse settings. By analyzing a wide range of interventions and their outcomes, it demonstrates the effectiveness of various intervention approaches. Additionally, this study identifies trends and patterns across interventions, highlighting which strategies are most successful in reducing treatment abandonment and improving patient outcomes.

Implications of all the available evidence: The collective evidence underscores the importance of implementing multifaceted interventions to address the complex factors contributing to treatment abandonment. Strategies that combine socioeconomic support, clinical care improvements, and psychosocial interventions show promising results in reducing abandonment rates and improving patient outcomes. Patient tracking systems emerged as important components of interventions across categories. Policymakers, healthcare providers, and stakeholders can use these findings to inform the development and implementation of targeted interventions tailored to specific contexts and patient populations. Furthermore, this study highlights the need for improved research methodologies and refinement of intervention strategies to optimize their impact on reducing treatment abandonment and improving patient care.

