## Appendix A for "Interventions to Reduce Pediatric Cancer Treatment Abandonment in Low- and Middle-Income Countries: A Scoping Review"

**Link to World Bank list:**

**https://datahelpdesk.worldbank.org/knowledgebase/articles/906519-world-bank-country-and-lending-groups**

**Medline Ovid search strategy**

1.	Developing Countries/

2.	(("low* income*" or "middle* income*" or "low-and-middle income" or "low-middle income*" or "low or middle income*" or "upper-middle income*" or developing or underdevelop* or "under-develop*" or "less-develop*" or "least-develop*" or "resource* constrain*" or "constrain* resource*" or "resource* limit*" or "limit* resource*" or impoverish* or poverty* or poor*) adj3 (countr* or nation* or state* or region* or world* or economy* or economies* or setting* or population*)).ti,ab,kw,kf.

3.	(LMIC* or LAMIC* or "third-world*" or "3rd world*").ti,ab,kw,kf.

4.	exp Afghanistan/ or exp Albania/ or exp Algeria/ or exp Angola/ or exp Argentina/ or exp Armenia/ or exp Azerbaijan/ or exp Bangladesh/ or exp Belarus/ or exp Belize/ or exp Benin/ or exp Bhutan/ or exp Bolivia/ or exp "Bosnia and Herzegovina"/ or exp Botswana/ or exp Brazil/ or exp Bulgaria/ or exp Burkina Faso/ or exp Burundi/ or exp Cabo Verde/ or exp Cambodia/ or exp Cameroon/ or exp Central African Republic/ or exp Chad/ or exp China/ or exp Colombia/ or exp Comoros/ or exp "Democratic Republic of the Congo"/ or exp Congo/ or exp Costa Rica/ or exp Cote d'Ivoire/ or exp Cuba/ or exp Djibouti/ or exp Dominica/ or exp Dominican Republic/ or exp Ecuador/ or exp Egypt/ or exp El Salvador/ or exp Equatorial Guinea/ or exp Eritrea/ or exp Eswatini/ or exp Ethiopia/ or exp Fiji/ or exp Gabon/ or exp Gambia/ or exp "Georgia (Republic)"/ or exp Ghana/ or exp Grenada/ or exp Guatemala/ or exp Guinea/ or exp Guinea-Bissau/ or exp Guyana/ or exp Haiti/ or exp Honduras/

5.	exp India/ or exp Indonesia/ or exp Iran/ or exp Iraq/ or exp Jamaica/ or exp Jordan/ or exp Kazakhstan/ or exp Kenya/ or exp Kiribati/ or exp "Democratic People's Republic of Korea"/ or exp Kosovo/ or exp Kyrgyzstan/ or exp Laos/ or exp Lebanon/ or exp Lesotho/ or exp Liberia/ or exp Libya/ or exp "Republic of North Macedonia"/ or exp Madagascar/ or exp Malawi/ or exp Malaysia/ or exp Maldives/ or exp Mali/ or exp Marshall Islands/ or exp Mauritania/ or exp Mauritius/ or exp Mexico/ or exp Micronesia/ or exp Moldova/ or exp Mongolia/ or exp Montenegro/ or exp Morocco/ or exp Mozambique/ or exp Myanmar/ or exp Namibia/ or exp Nauru/ or exp Nepal/ or exp Nicaragua/ or exp Niger/ or exp Nigeria/ or exp Pakistan/ or exp Palau/ or exp Papua New Guinea/ or exp Paraguay/ or exp Peru/ or exp Philippines/

6.	exp Romania/ or exp Russia/ or exp Rwanda/ or exp Samoa/ or exp "Sao Tome and Principe"/ or exp Senegal/ or exp Serbia/ or exp Seychelles/ or exp Sierra Leone/ or exp Solomon Islands/ or exp Somalia/ or exp South Africa/ or exp South Sudan/ or exp Sri Lanka/ or exp Saint Lucia/ or exp "Saint Vincent and the Grenadines"/ or exp Sudan/ or exp Suriname/ or exp Syria/ or exp Tajikistan/ or exp Tanzania/ or exp Thailand/ or exp Timor-Leste/ or exp Togo/ or exp Tonga/ or exp Tunisia/ or exp Turkey/ or exp Turkmenistan/ or exp Tuvalu/ or exp Uganda/ or exp Ukraine/ or exp Uzbekistan/ or exp Vanuatu/ or exp Venezuela/ or exp Vietnam/ or exp Yemen/ or exp Zambia/ or exp Zimbabwe/

7.	(Afghanistan* or Albania* or Algeria* or Angola* or Argentina* or Armenia* or Azerbaijan* or Bangladesh* or Belarus* or Belize* or Benin* or Bhutan* or Bolivia* or Bosnia* or Herzegovina* or "Bosna i Her*" or Botswana* or Brazil* or Bulgaria* or "Burkina Faso*" or Burundi* or "Cabo Verde*" or "Cape Verde*" or Cambodia* or Cameroon* or "Central African Republic*" or Chad* or China* or Colombia* or Comoros* or Congo* or "Costa Rica*" or "Cote d'Ivoire*" or "Cote d Ivoire*" or "Cote dIvoire*" or "Ivory Coast*" or Cuba* or Djibouti* or Dominica* or "Dominican Republic*" or Ecuador* or Egypt* or "El Salvador*" or ElSalvador* or "Equatorial Guinea*" or Eritrea* or Eswatini* or Ethiopia* or Fiji* or Gabon* or Gambia* or Gaza* or Georgia* or Ghana* or Grenada* or Guatemala* or Guinea* or "Guinea-Bissau*" or Guyana* or Haiti* or Honduras*).ti,ab,kw,kf.

8.	(India* or Indonesia* or Iran* or Iraq* or Jamaica* or Jordan* or Kazakhstan* or Kenya* or Kiribati* or "Democratic People's Republic of Korea*" or "North Korea*" or Kosovo* or Kyrgyzstan* or "Kyrgyz* Republic*" or Laos* or "Lao People's Democratic Republic*" or "Lao PDR*" or Lebanon* or Lesotho* or Liberia* or Libya* or Macedonia* or Madagascar* or Malawi* or Malaysia* or Maldives* or Mali* or "Marshall Island*" or Mauritania* or Mauritius* or Mexico* or Mejico* or Micronesia* or "Micro-nesia*" or Moldova* or Mongolia* or Montenegro* or Morocco* or Mozambique* or Myanmar* or Namibia* or Nauru* or Nepal* or Nicaragua* or Niger* or Pakistan* or Palau* or "Papua New Guinea*" or Paraguay* or Peru* or Philippine*).ti,ab,kw,kf.

9.	(Romania* or Russia* or Rwanda* or Samoa* or "Sao Tome*" or Principe* or Senegal* or Serbia* or Seychelles* or Siberia* or "Sierra Leone*" or "Solomon Islands*" or Somalia* or "South Africa*" or Sudan* or "Sri Lanka*" or "Saint Lucia*" or "St Lucia*" or "Saint Vincent*" or "St Vincent*" or Grenadines* or Suriname* or Syria* or Tajikistan* or Tanzania* or Thailand* or Timor* or Togo* or Tonga* or Tunisia* or Turkey* or Turkiye* or Turkmenistan* or Tuvalu* or Uganda* or Ukraine* or Uzbekistan* or Vanuatu* or Venezuela* or Vietnam* or "West Bank*" or Yemen* or Yugoslav* or Zambia* or Zimbabwe*).ti,ab,kw,kf.

10.	exp Africa/ or exp Caribbean Region/ or Central America/ or Latin America/ or exp South America/

11.	(Africa* or Caribbean* or caribean* or carribbean* or carribean* or "West Indies*" or "Central America*" or "Latin America*" or "South America*" or subsaharan* or "sub-saharan*" or "south of the sahara*").ti,ab,kw,kf.

12.	or/1-11

13.	exp Pediatrics/ or exp Infant/ or exp Child/ or Adolescent/

14.	(pediatric* or paedatric* or infan* or newborn* or "new-born*" or neonat* or "neo-nat*" or baby* or babies* or toddler* or child* or adolescen* or tween* or teen* or juvenile* or youth* or AYA or AYAs or preschool* or "pre-school*" or "nursery school*" or kindergarten* or "kinder-garten*" or kindergarden* or "kinder-garden*" or schoolchild* or "school-age*" or "middle school" or "junior high*" or "high school*").ti,ab,kw,kf,ja,jn,jw,nj,nw.

15.	or/13-14

16.	exp Neoplasms/

17.	(neoplas* or "neo-plas*" or cancer* or blastom* or carcino* or gliom* or leukemia* or lymphoma* or malignan* or melanom* or neuroblastom* or oncolog* or osterosarcom* or retinoblastom* or sarcom* or tumor* or tumour*).ti,ab,kw,kf,ja,jn,jw,nj,nw.

18.	or/16-17

19.	Treatment Refusal/ or "Treatment Adherence and Compliance"/ or Patient Compliance*/ or Patient Dropouts/ or No-Show Patients/

20.	((cancer* or treatment* or therap* or care* or regimen* or patient*) adj4 (abandon* or abscond* or elope* or refus* or stop* or cease* or cessation or adher* or comply* or complie* or complian* or noncomplian* or absent* or absence* or dropout* or "drop-out*" or "no-show*" or attendance*)).ti,ab,kw,kf.

21.	or/19-20

22.	Internet-Based Intervention/ or Clinical Protocols/

23.	(interven* or "inter-ven*" or protocol*).ti,ab,kw,kf.

24.	Patient Education Handout/ or Patient Education as Topic/ or Pamphlets/

25.	(brochure* or educat* or handout* or "hand-out*" or pamphlet*).ti,ab,kw,kf.

26.	exp Counseling/ or Self-Help Groups/

27.	(counsel* or "self-help*" or "support* group*").ti,ab,kw,kf.

28.	exp Financial Support/

29.	((accommodat* or economic* or financ* or hotel* or housing* or incentiv* or money* or monetary* or fund*) adj3 (aid* or assist* or educat* or empower* or endow* or help* or inclusion* or literacy* or provid* or provis* or support* or subsid*)).ti,ab,kw,kf.

30.	Patient Advocacy/ or Patient Navigation/ or Patient Identification Systems/ or Health Smart Cards/ or Radio Frequency Identification Device/ or Monitoring, Ambulatory/

31.	(patient* adj3 (advoca* or empower* or ID or identification* or navigat* or RFID or RFIDs or monitor* or track* or surveil*)).ti,ab,kw,kf.

32.	((home* or ambulatory*) adj3 (monitor* or track* or surveil*)).ti,ab,kw,kf.

33.	(smartcard* or "smart card*").ti,ab,kw,kf.

34.	"twinning program*".ti,ab,kw,kf.

35.	Cell Phone/ or Smartphone/ or Text Messaging/ or Electronic Mail/ or Mobile Applications/

36.	(cellphone* or "cell* phone*" or email* or "e-mail*" or "instant messag*" or "mobile app*" or "mobile device*" or "mobile phone*" or "phone app*" or smartphone* or "smart-phone*" or text*).ti,ab,kw,kf.

37.	((barrier* or hinder* or hindrance* or obstacle* or "unmet need*") adj3 (address* or answer* or eliminat* or fulfill* or minimiz* or minimis* or overcom* or "over-com*" or reduc* or solv* or solution*)).ti,ab,kw,kf.

38.	or/22-37

39.	12 and 15 and 18 and 21 and 38
